# *G6PD* variant distribution in sub-Saharan Africa and potential risks of using chloroquine/hydroxychloroquine based treatments for COVID-19

**DOI:** 10.1101/2020.05.27.20114066

**Authors:** Jorge da Rocha, Houcemeddine Othman, Caroline T. Tiemessen, Gerrit Botha, Michèle Ramsay, Collen Masimirembwa, Clement Adebamowo, Ananyo Choudhury, Jean-Tristan Brandenburg, Mogomotsi Matshaba, Gustave Simo, Francisco-Javier Gamo, Scott Hazelhurst, as members of the H3Africa Consortium

## Abstract

Chloroquine/hydroxychloroquine have been proposed as potential treatments for COVID-19. These drugs have warning labels for use in individuals with glucose-6-phosphate dehydrogenase (G6PD) deficiency. Analysis of whole-genome sequence data of 458 individuals from sub-Saharan Africa showed significant *G6PD* variation across the continent. We identified nine variants, of which four are potentially deleterious to G6PD function, and one (rs1050828) that is known to cause G6PD deficiency. We supplemented data for the rs1050828 variant with genotype array data from over 11,000 Africans. Although this variant is common in Africans overall, large allele frequency differences exist between sub-populations. African sub-populations in the same country can show significant differences in allele frequency (e.g. 16.0% in Tsonga vs 0.8% in Xhosa, both in South Africa, *p* = 2.4 × 10^−3^). The high prevalence of variants in the *G6PD* gene found in this analysis suggests that it may be a significant interaction factor in clinical trials of chloroquine and hydrochloroquine for treatment of COVID-19 in Africans.

## 1 Introduction

Chloroquine and hydroxychloroquine (CQ/HCQ) are currently undergoing clinical trials as treatments for Coronavirus disease 2019 (COVID-19) which is caused by the severe acute respiratory syndrome coronavirus 2 – SARS-Cov-2 [32]. CQ/HCQ and other aminoquinolines have pharmacogenomic associations with the glucose-6-phosphate dehydrogenase (*G6PD*) gene [23]. Aminoquinolines are suspected to exert their antimalarial effect by increasing oxidative stress via production of haem-based reactive oxygen species [16]. The G6PD enzyme is responsible for the production of nicotinamide adenine dinucleotide phosphate (NADPH) which is required in the glutathione mediated detoxification of reactive oxygen species [21]. In the case of inactive/deficient G6PD, the NADPH supply may not be sufficient to neutralize the reactive oxygen species induced by CQ/HCQ and other drugs with similar mechanisms of action.

G6PD deficiency is common globally, particularly in African populations (14% of males) [27]. Individuals with the deficiency are at risk for haemolytic anaemia which can be triggered by infections, certain foods or medications. G6PD deficiency is an X-linked disorder. It mostly occurs in males who are hemizygous for deleterious variants of the *G6PD* gene and in females with homozygous deleterious variants. Symptoms have also been observed in females with heterozygous combinations due to X-inactivation effects [40]. Three common haplotype arrangements have been defined for the gene, the B (wild type), A and A-(deficiency). The *G6PD* A ‘haplotype’ is denoted by the rs1050829 variant (c.376A>G). The rs1050829 C variant is not linked to decreased G6PD activity and occurs in 10 - 30% of sub-Saharan Africans [38, 19].

The A– haplotype (which is associated with G6PD enzyme deficiency) is formed by a combination of two variants, one of which is a deleterious variant, while the other is the rs1050829 C variant. A commonly observed combination is that of rs1050828 T (c.202G>A), which occurs in 10% of sub-saharan Africans, and rs1050829 [23]. The *G6PD* A– haplotype causes between 10-60% reduction in G6PD enzyme activity. The A-*G6PD* haplotype is classified as a World Health Organisation (WHO) Class III variant [1, 23]. Strong linkage disequilibrium exists between the rs1050828 T and rs1050829 C variants [39]. As the rs1050829 C allele is more common, it is likely that that rs1050828 T emerged after rs1050829 C, and then increased in frequency due to positive selection in Africans [33]. As rs1050829 C has no effect on G6PD deficiency, it is reasonable to report G6PD deficiency based on rs1050828 genotype combinations alone.

The FDA has issued warnings on the use of CQ and HCQ in G6PD deficient individuals due to high risk of haemolytic anaemia, although these are not contraindicated [36, 37]. Acute haemolytic effects following HQC treatment for COVID-19 have been reported in a single male case [8] who is suspected of carrying the *G6PD* Mediterranean variant. Whereas CQ is not known to induce severe haemolytic effects when used as an antimalarial in G6PD deficient individuals, in contrast to primaquine [34] or chloroproguanil [9], the risk of its therapeutic use in G6PD deficient, COVID-19 patients is unknown.

In this paper, we evaluate the prevalence of variants in G6PD gene in individuals of African ancestry. We suggest that variations in the *G6PD* gene could significantly affect risk of adverse effects of CQ/HCQ, and recommend that this should be evaluated in clinical trials of CQ/HCQ treatment for COVID-19. We also report the prevalence of a key *G6PD* variant, rs1050828, in 11,030 Africans from four countries in west, east and southern Africa, and show that not only is the variant allele common in Africa overall, but that there are very large differences between different groups, even between those who reside in close proximity.

## 2 Methods

The dataset used was assembled as a collaborative project of the Human Heredity and Health in Africa (H3A) Consortium. The high coverage African ADME Dataset (HAAD) was sourced from H3A, other African collaborations and the Simon Foundation’s Genome Diversity Project [18]. HAAD consists of high-coverage sequences from 458 Sub-Saharan African individuals from 15 countries, with 8 of these countries contributing data from more than 25 individuals (Nigeria, Ghana, Burkina Faso, Cameroon, Benin, Botswana, Zambia, and South Africa) (Figure 1 A). HAAD BAMS were aligned to GRCh37 with bwa-mem v0.7.10-v0.7.17 [14]. Variants were called with Haplotype-Caller in gVCF mode using GATK v.4.0.8.1 HAAD gVCFs (along with gVCFs produced with African 1000 Genomes Project data (KGA) [2]) were combined with GATK’s CombineGVCF (v.4.0.8.1), and jointly called with GenotypeGVCFs (v4.1.3.0) and followed GATK’s best practice guidelines. VQSR was used to select high quality sites with PASS ratings. All related workflows for data preparation can be found at https://github. com/h3abionet/recalling. The *G6PD* canonical gene region (chrX:153759606-153775469) was extracted with bcftools v1.9, and variants were annotated (e.g. as missense, intronic etc.) with variant effect predictor (VEP) v92.0 [24] and SNPeff v4.3t [6]. Coding variants were selected for analysis if they meet a *QUAL >* 50 quality score. Functional annotation for these variants was performed using dbNSFP [15] to retrieve scores for five predictive toolsets (LRT, MutationAssessor, PROVEAN, VEST3 and CADD), which were then used for prediction based on a pharmacogene optimised model [43]. These form part of the “g_miner” workflow which is available at: https://github.com/hothman/PGx-Tools/tree/master/workflows/g_miner. PLINK [4] was used to call allele frequencies in every country in the HAAD dataset. Statistical analyses were conducted using R v3.63 [28]. The test for equal or given proportions was used to calculate allele frequency confidence intervals (CI) at the 95% significance level. Fisher’s exact test was used to assess significant differences in allele frequency between two populations, at the 5% significance threshold.

**Figure 1:**
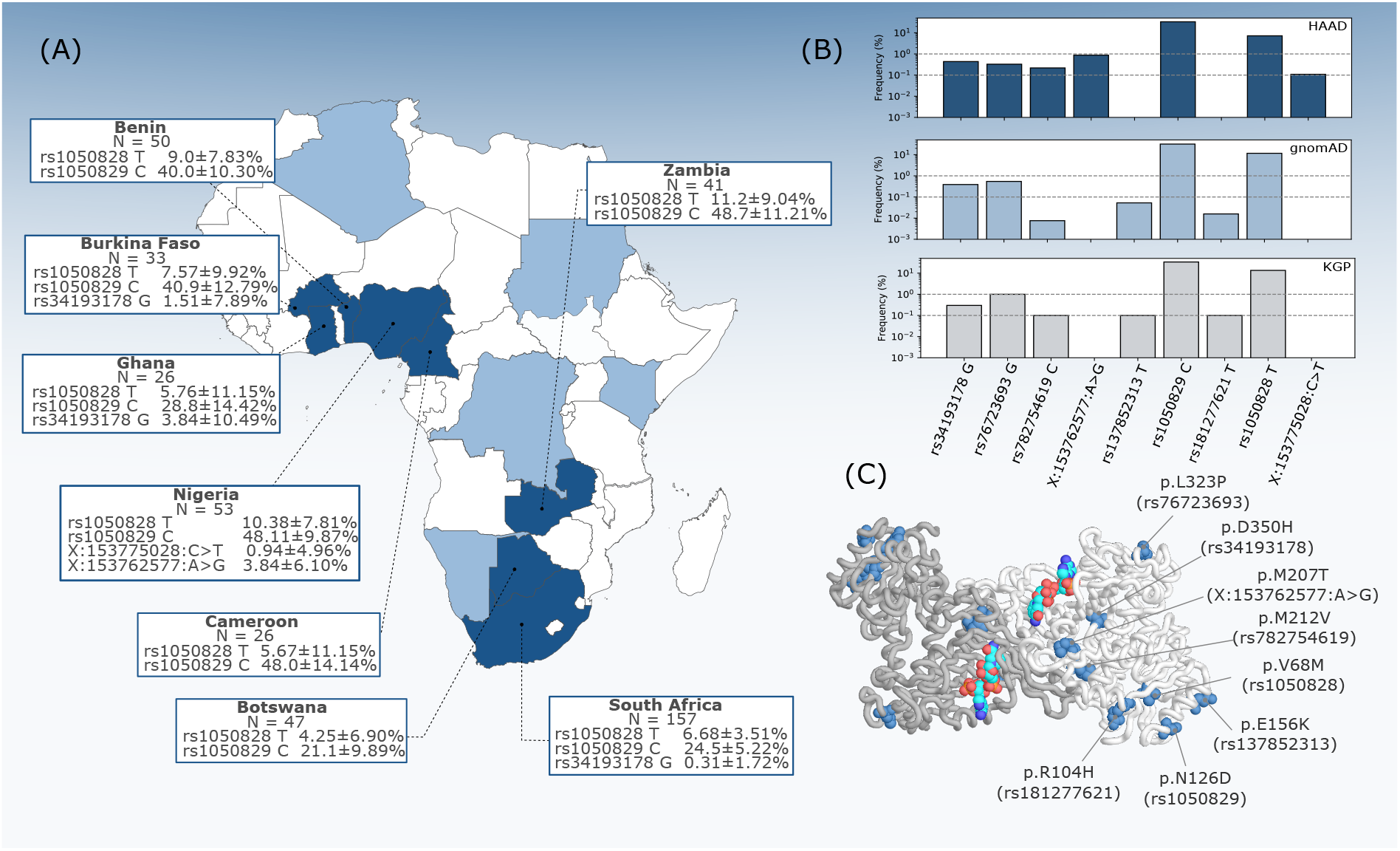
*G6PD* missense variant distribution across African populations. (A) G6PD allele frequencies in populations from HAAD countries. Confidence intervals for allele frequencies based on the equal or given proportions test the 95% significance level. (B) Allele frequencies of missense variants in HAAD, and African superpopulation groups from gnomAD and the KGP. (C) Structural representation of the G6PD homodimer with missense residues highlighted in blue color on both chains with bound NADP (NADP shown in red-turquoise-blue).

The impact of the variant on the protein structure was assessed by DynaMut [31] using the crystal structure of Canton G6PD modified to the wild type form [12]. Stability related metrics calculated by Dynamut include the change in Gibbs free energy (∆∆*G*).

The A-haplotype in this study is defined by the presence of the rs1050828 T allele (alleles assessed in forward orientation, this corresponds to the c.202G>A nomenclature for cDNA NM_001042351.2). The presence of rs1050829 C (corresponding to c.376A>G) is assumed due to strong linkage disequilibrium with rs1050828 T [39]. The rs1050828 genotypes were previously generated by the AWI-Gen Project [30] on 11,062 sub-Saharan Africans from 6 sites in Ghana, Burkina Faso, Kenya and South Africa using the H3A Custom Genotyping Array (https://www.h3abionet.org/h3africa-chip). PLINK [4] was used to remove any individuals with more than 1% missingness or genotypes which conflicted with declared sex. This left 11,030 individuals in all. The cluster plots of the genotype calls (male, female, all) are consistent with a well-genotyped single nucleotide polymorphism on the X chromosome. Minor allele frequency (MAF) was computed for the overall AWI-Gen data set as well as various sub-groups, as determined by self-identified ethnicity. The data was analysed using a custom Python script using the pandas-plink library (https://github.com/limix/pandas-plink).

## 3 Results

### 3.1 Variation from high coverage data

Nine coding (missense) variants were identified in the WGS, of which seven have been previously described, and have rsIDs allocated on dbSNP (nomenclature referred to by rsID throughout, as defined in Table 1). No loss of function type variants were detected.

**Table 1:** *G6PD* missense variants detected within HAAD and KGA population datasets and their relative stability effect (ΔΔG) for G6PD protein.

| Variant ID | Nucleotide | Amino Acid | $\Delta\Delta G$ (kcal/mol) |
| --- | --- | --- | --- |
| rs34193178 | 1048G>A | D350H | 0.374 |
| rs76723693 | 968T>C | L323P | -0.872 |
| rs782754619 | 634A>G | M212V | -1.147 |
| rs181277621 | 311G>A | R104H | -1.401 |
| chrX:g.153762577A>G | T620C | M207T | -1.145 |
| chrX:g.153775028C>T | G58A | N/A | N/A |
| rs137852313 | 466G>A | E156K | 0.539 |
| rs1050829 | 376A>G | N126D | 0.887 |
| rs1050828 | 202G>A | V68M | -1.347 |

Nucleotide positions based on cDNA NM\_001042351.2 cDNA (chrX:g.153775028C>T G58A is indicated by NM\_000402.4). Amino acid positions based on the protein sequence of from 6E07 PDB structure.

Figure 1B shows the distribution of the *G6PD* missense variants across African populations, along with comparisons to their overall frequency in the African data from the Genome Aggregation Database (gnomAD) [13] and the 1000 Genomes Project (KGP) [2]. Of these nine variants, seven have at least one prediction score from the pharmacogene model indicating deleterious impact. The variants rs1050829 (chrX:g.153763492T>C), and chrX:g.153775028C>T had no predictive score reaching model cutoff criteria.

G6PD deficiency (A–), as defined by the rs1050828 T allele, is widely distributed across the African continent (Figure 1 A). There are notable differences in frequency across different groups, and we note that frequency does not necessarily correlate with the geographic location of the country. The highest frequency observed in HAAD populations was in Zambians (11.2±9.04%, 95% CI), whereas the lowest was in Batswana (4.25±6.90%, 95% CI) their geographic neighbours. At the low sample number, this difference is not significant (p = 0.09115), and the confidence interval is large, however, we note that allele frequency is not uniform across these and other HAAD African groups.

Two other missense variants in HAAD, rs76723693 (chrX:g.153761240A>G) and rs34193178 (chrX:g.153761160C>G), have robust predictions as being functionally deleterious (all model tools in consensus). rs76723693 is defined as part of the A-haplotype and although very rare in Africans, it has been characterised in other global populations [23]. The rs34193178 variant is rare overall, but is found at a 1.5±7.89% in HAAD Burkina Faso and 3.84±10.49% in HAAD Ghana populations (95% CI). It is also present in HAAD South Africans at 0.3±1.72% (95% CI).

The two variants chrX:g.153762577A>G and chrX:g.153775028C>T were not found in dbSNP151, and have not been reported in the gnomAD and the KGP databases (Figure 1B). The chrX:g.153762577A>G variant is only found in the HAAD Nigerian population, at 3.8±6.10% (95% CI), but not seen in the KGA Nigerian Esan or Yoruba populations. The chrX:g.153775028C>T is a singleton variant, and is only present in pre-protein structures and is thus not displayed in Figure 1C.

Four of the variants (rs1050828, rs137852313, rs1050829, rs181277621) are located in the co-enzyme domain; rs782754619 and chrX:g.153762577A>G belong to the *α* + *β* domain of G6PD buried in the protein core; and rs76723693 and rs34193178 are exposed to the solvent. All the corresponding amino acid residues (with the exception of rs137852313) are either densely packed against other residues within the structure of G6PD, or they establish polar contacts that appear to stabilize local conformations of nearby segments. Structural predictions (as based on ΔΔG (kcal/mol) -Table 1) of protein variant effect show a destabilizing effect for rs76723693, rs782754619, chrX:g.153762577A>G, rs181277621, and rs1050828. The rs34193178, rs137852313 and rs1050829 variants showed a stabilizing effect.

### 3.2 High variability of rs1050828 allele frequency in Africa

Previous studies have shown that the MAF of rs1050828 is relatively high in African populations [20, 7]. We further show that it is also extremely variable - even within the same geographical region. Table 2 shows the minor allele frequency in the AWI-Gen study (over 11,030 participants). A full per-group analysis is not possible in this rapid communication. However, we show the MAF in selected groups with at least 180 individuals.

**Table 2:** Minor allele frequency (MAF) of rs1050828 (T) in selected groups from the AWI-Gen study genotype data. Het: Proportion of females heterozygous (%), Hom: Proportion of females homozygous for the alternate allele (%). Note that 100 of the HAAD SA individuals are included in this genotyping study (< 2% of the samples)

| Group | All |  | Males<br>MAF | Females |  |  |  |
| --- | --- | --- | --- | --- | --- | --- | --- |
|  | N | MAF |  | N | MAF | Het | Hom |
| All Genotyped Samples | 11,030 | 12.6 | 11.9 | 6033 | 12.8 | 21.3 | 2.2 |
| South Africa |  |  |  |  |  |  |  |
| Tsonga | 2,132 | 16.0 | 14.1 | 1209 | 16.7 | 26.9 | 3.2 |
| BaPedi-Tswana-Sotho | 1,849 | 5.5 | 5.1 | 1233 | 5.6 | 10.5 | 0.4 |
| Xhosa | 180 | 0.8 | 0.9 | 68 | 0.7 | 1.5 | 0.0 |
| Ghana and Burkina Faso |  |  |  |  |  |  |  |
| Mampruga-Mossi-Gouronsi-Kassena-Nankana | 3,723 | 18.6 | 17.6 | 1,888 | 19.1 | 30.7 | 3.7 |
| Kenya |  |  |  |  |  |  |  |
| Luhya-Luo-Kamba | 978 | 11.0 | 8.6 | 454 | 12.3 | 20.7 | 2.0 |
| Kikuyu | 655 | 6.8 | 6.3 | 434 | 6.9 | 12.4 | 0.7 |

The rs1050828 T variant frequency in 11,030 individuals was 12.6%, close to the values reported in gnomAD (11.6%) and KGP (13.5%). Overall, 11.9% of African males carried the variant on their X chromosome while 2.2% of females were homozygous for the T allele. A moderate *G6PD* deficiency (10-60% residual enzyme activity) (WHO Class III) is likely to be present in individuals with such genotypes [1]. Although rare, the deficiency can present in heterozygous females depending on X-inactivation effects [40]. There was a significant difference in the variant frequency among self-identified ethnic groups in South Africa and Kenya. The frequency among the Tsonga was 16.0% which is substantially different from 0.8% found in the Xhosa (*p* = 2.4 × 10^−3^) and 5.5% in the BaPedi-Tswana-Sotho (*p* = 2.2 × 10^−16^) ethnolinguistic groups. The T variant appears at markedly different frequency in different Kenyan groups - 6.8% in the Kikuyu and 11% in the combined Luhya-Luo-Kamba groups (*p* = 4.3 × 10^−3^).

We tested for deviation from Hardy-Weinberg equilibrium (HWE) in the females in each of the groups except for the Xhosa (in which which there was only 1 person who had the variant allele) and no significant deviation could be shown (lowest p-value was 0.38). In women overall the expected proportion of women who are heterozygous under assumption of HWE is 0.224 while the observed proportion was 0.213. While this is highly significant (p = 2.7 × 10^−4^), this deviation is not unexpected given the very significant differences in MAF between groups.

## Discussion

Current clinical studies that have used CQ/HCQ for treatment of COVID-19 have not explicitly taken into account the potential risks posed by G6PD deficiency [3, 5, 10]. G6PD deficiency is known to be common in Africans. In the present study, we assessed *G6PD* gene variation in African populations, and noted the high prevalence of a common deleterious allele - rs1050828. Although common, there are large differences in frequency for this variant, even between populations that are geographic neighbours. The differences may be explained by selective pressures in regions where malaria is/was common [11], as G6PD deficiency may convey resistance to malaria [22]. Such differences have been previously reported to occur even within countries, as in Botswana, where a decreasing trend in frequency of this variant occurs from the north-west to south-east [26]. Another study in South Africans (n = 181) from the Mpumalanga province reported the allele frequency of A-to be 14% [29], which is similar to findings from AWI-Gen genotype data, where a MAF of 16% was found in Tsonga-speaking individuals. However, other SA groups, such as the Xhosa in particular (MAF 0.8%) had much lower frequencies. This highlights the limitation of reporting allele frequency by country rather than ethnolinguistic groups. African populations undergoing COVID-19 CQ/HCQ treatment trials may not have the same relative frequency of this allele as others. Thus we urge that models for *G6PD* related effects based on a single proxy African population are not directly transferable to other Africans, even those close geographically.

We observed other potentially deleterious variants in the African populations we studied. For instance, rs34193178 and chrX:g.153762577A>G, which were not found across all populations, do not have well characterised effects, and are unlikely to be included in assays to type well known *G6PD* variants. If these have a functional impact on G6DPD, they may add complexities to studies assessing the presence of rs1050828 and rs1050829 alone. The chrX:g.153762577A>G variant in particular has structural evidence for deleterious functional impact.

Although use of either CQ/HCQ is not new in African populations, the dosage and duration of CQ/HCQ treatment for COVID-19 may lead to higher prevalence of adverse effects related to G6PD deficiency. Acute haemolysis has been observed in a G6PD deficient male suspected of carrying the *G6PD* Mediterranean variant (rs5030868) who was treated with lopinavir and HCQ [8]. Although the Mediterranean variant is known to induce a greater sensitivity to pro-oxidant drugs than the A-variant [43], the observation of haemolytic effects highlights the relevance of assessing the impact of G6PD deficiency on CQ/HCQ that is repurposed for COVID-19 treatment. Non-haemolytic adverse effects have also been noted in other recent trials. A CQ trial in Brazilians noted severe adverse reactions related to QT elongation [3], and the high doses (600 mg twice daily) may pose greater risks for individuals with G6PD deficiency. The proportion of African admixture for the patients assessed in this study was not disclosed. A recent trial in U.S. Veterans showed increased risk of mortality in patients treated with HQC [17]. In addition, it is currently unknown how G6PD deficiency may affect COVID-19 disease progression. G6PD deficient cells have been found to be more vulnerable to human *alphacoronavirus* 229E infection in vitro, which correlated with elevated oxidant production [41], although it is not yet known if this effect would also be seen with the novel SARS-Cov-2 virus. Monitoring of G6PD deficiency throughout COVID-19 trials and studies in Africans may therefore also reveal other factors which are not limited only to effects of drug response. Such studies could make use of a rapid enzymatic assay for G6PD deficiency in lieu of DNA-based assays. These are available, though it should be noted that their sensitivity is lower in females [42].

As a final note, the applicability and evidence basis of CQ/HCQ as COVID-19 treatments have recently been well reviewed [35, 25] and their current implementation have been questioned [35]. These reviews conclude that there is currently insufficient evidence for the use of CQ/HCQ as COVID-19 treatments. As trials are still ongoing, we urge the consideration of G6PD deficiency related effects in African populations participating in these studies.

## 4 Conclusion

Given our findings of the large heterogeneity of the *G6PD* gene, variants associated with G6PD deficiency in Sub-Saharan Africa, and the possible presence of other uncharacterised deleterious variants, it is important to consider the potential impact of these variants before widespread use of CQ/HCQ as COVID-19 treatments for African populations. Distinct African ethnolinguistic groups can have vastly different frequencies of G6PD deficiency, thus clinical studies of CQ/HCQ for COVID-19 in Africans should be conducted on diverse African populations, and with monitoring for haemolysis and/or anaemia. Targeted sequencing of *G6PD* among study participants would provide important insights into the risks of adverse effects at therapeutic doses which might lead to dosage adjustment.

## Data Availability

Datasets used in this study were generated as part of the Human Heredity and Health in Africa Project (H3Africa). H3Africa is committed submit these datasets to the European Genome-Phenome Archive (EGA), where access can be obtained through the H3Africa Data and Biospecimen Access Committee.

https://ega-archive.org/search-results.php?query=h3africa

## Acknowledgements

This paper uses the results of data and analysis from the GSK/H3A ADME project. We acknowledge the generous collaboration of the H3A AWI-Gen, CAfGEN, and TrypanoGEN groups, the support of the Cell Biology Research Lab, NICD/Wits, the Awadalla Lab at the Université de Montréal, Gustave Simo from the University of Dschang, Clement Adebamowo at the Institute for Human Virology, Abuja, Martin Simuunza from the Department of Disease Control at the Univeristy of Zambia, and Gabriel Anabawi from the University of Botswana. We sincerely thank the data providers, and study participants of these groups for their contribution. The AWI-Gen Collaborative Centre is funded by the NIH/NHGRI (Grant U54HG006938) as part of the H3Africa Consortium. MR is a South African Research Chair in Genomics and Bioinformatics of African Populations hosted by the University of the Witwatersrand, funded by the Department of Science and Technology, and administered by National Research Foundation of South Africa (NRF). The TrypanoGEN project was funded by the Wellcome Trust, study number 099310/Z/12/Z. The Collaborative African Genetics Network (CAfGEN) is funded by the NIH/NHGRI (grant CAfGEN 1U54AI110398). The whole genome sequencing of the H3A Data was supported by a grant from the National Human Genome Research Institute, National Institutes of Health (NIH/NHGRI) U54HG003273. Cell Biology Research Lab component: This work is based on the research supported by grants awards from the Strategic Health Innovation Partnerships (SHIP) Unit of the South African Medical Research Council, a grantee of the Bill & Melinda Gates Foundation, and the South African Research Chairs Initiative of the Department of Science and Technology and National Research Foundation of South Africa (84177). JdR was partially funded by the SA National Research Foundation (SFH170626244782). GSK had no part in the design of this study or analysis presented here and the views and opinions expressed are not necessarily those of GSK. The authors acknowledge the financial support of GSK.

## Notes

### Competing Interest Statement

The authors have declared no competing interest.

### Author Declarations

Human Research Ethics Committee (Medical), University of the Witwatersrand; Clearance Certificate NO. M190541

